# An AI-Powered Smartphone Application for Universal and Standardized Reading and Interpretation of Lateral Flow Assays

**DOI:** 10.64898/2026.05.14.26352875

**Authors:** David Bermejo-Peláez, Oscar Darias, Lucia Pastor, Ramón Vallés, Nuria Diez, Lin Lin, Jaime García-Villena, Daniel Cuadrado, Alexander Vladimirov, Elisa Álamo, Maria Postigo, Mario Rodriguez-Dominguez, Rafael Cantón, Juan Luis Rodriguez-Tudela, Ana Alastruey Izquierdo, Andrea Marchiol, Maryi Lorena Segura Alba, Liliana Jazmín Cortés Cortés, Laura C. Bohorquez, Jose Miguel Rubio, Elena Dacal, Miguel Luengo-Oroz

## Abstract

**Introduction:** Lateral flow assays (LFAs) are indispensable rapid diagnostic tools in healthcare, enabling point-of-care diagnosis critical for patient management and support disease burden assessment and surveillance when results are properly recorded. However, misinterpretation errors and unreported cases remain a concern. A quality-assured, affordable Ai-powered tool, supporting the decision-making during result interpretation could promote proper disease monitoring and epidemiological surveillance. Here, we describe the performance of a universal AI model to digitize and interpret results from multiple LFA types through a smartphone application, a step that could ultimately enable standardized and digitally reportable test outcomes.

**Methods:** The AI algorithm was evaluated in 17 LFA types, including both 2-band and 3-band tests for different diseases and manufacturers. The model was trained on a dataset of 22,576 images captured under diverse lighting conditions with different smartphone models and using a custom mobile application, TiraSpot (Spotlab, Madrid, Spain). To assess generalizability, a leave-one-out cross-validation was applied, wherein each LFA type was iteratively excluded from training and used for testing. Model performance was evaluated using bootstrapping on the inference dataset.

**Results:** In the assessment of the model’s ability to generalize to new LFA types not previously analyzed (not included during development), the model achieved an overall AUC of 94.3% for second band detection. This overall performance was enhanced to 99.3% (Sensitivity=98,6%; Specificity=98%) after training with 50 images of each LFA type, highlighting the benefit of additional data for specific LFA types. For the third band detection, where less training data was available, the system achieved an overall AUC of 83.9% for unseen LFAs, improving to 94.2% (Sensitivity=92.9%; Specificity=87,9%) after training with 50 images of each LFA type.

**Conclusion:** This system demonstrates the feasibility of an AI-powered universal digital reader for interpreting LFA results from diverse test types using smartphone-captured images. Its compatibility with standard smartphones makes it a universal tool, enabling reliable LFA interpretation across devices and settings. By standardizing test interpretation and digitizing results, this tool could support decision making in result interpretation, enhancing epidemiological surveillance, particularly in resource-limited settings. Its adaptability across various infections highlights its potential to improve diagnostic consistency and support disease management in diverse healthcare settings.

## INTRODUCTION

Rapid Diagnostic Tests (RDTs), play a crucial role in patient management, treatment selection, and informing health systems in disease control and elimination strategies. Among RDTs, lateral flow assays (LFAs) constitute a significant subset, with hundreds of millions performed annually worldwide. These tests offer the potential of universal diagnosis and timely treatment initiation at the point-of-care (POC), decentralizing management of a wide variety of infectious diseases, as well as various non-communicable conditions. They offer a user-friendly and cost-effective approach to diagnostics, making it accessible for end users of different expertise levels and more affordable for health systems in resource-limited or decentralized settings *(1)*.

Despite these advantages, LFAs have several drawbacks, including potential user errors during test execution and results interpretation, as well as difficulties in traceability and records availability. Many of these challenges could be mitigated through a digital reporting system, however, the lack of an accessible electronic platform that supports the integration of LFAs results and patient clinical records into health systems hampers disease monitoring and data tracking (2–5). The paradigm of connected diagnostics is outlined under the REASSURED concept proposed by Land et al. *(6)*, including an additional requirement for real-time connectivity to the classic ASSURED diagnostic test criteria (affordable, sensitive, specific, user-friendly, rapid, equipment-free, delivered) *(7)*. This concept, also outlined in the WHO Target Product Profile (TPPs) for RDT readers *(8–10)*, emphasizes the critical need of digital connectivity across all diagnostic workflows to timely prompt treatment decisions, inform health systems on disease prevalence and allow for proper resources allocation and care management.

The current global availability and widespread use of smartphones have powerful implications for digital health care. Connection to mobile networks allows for real-time communication of test results, bridging gaps in patient care and epidemiological surveillance. Their connection with the Internet and cloud services makes them a highly valuable tool to develop software applications with artificial intelligence (AI) models able to automatically interpret the results of LFAs and facilitate immediate reporting of results. Several smartphone-based systems have demonstrated their capacity to automatically detect LFA results for different infections including HIV, SARS-CoV-2, *Plasmodium* and *Cryptococcus*, among others, performing both qualitative *(11–19)* and quantitative analyses *(20–24)*. These systems use different image processing approaches, from classical to more sophisticated machine learning or deep learning methods. However, most of these approaches rely on the use of additional hardware to be robust among different ambient lighting conditions or smartphone models and they are designed for the detection of a single analyte with specific LFA brands.

Few research studies have addressed the generalization capability of these digital platforms between different LFA types, brands and analytes *(12)*. In this paper, we describe an AI-based approach for universally reading and interpreting a wide range of LFAs, including dipstick formats without plastic cassettes. Central to the proposed approach is a smartphone application that proposes a protocol with simple instructions to standardize the digitization of LFA images under diverse field conditions without any external hardware. These images were then processed by a universal AI model, which accurately interpreted the results regardless of the LFA type. To facilitate broader data management and potential real-time surveillance, the system was also integrated with a cloud platform, allowing for aggregated visualization and refinement of results. By ensuring consistent, on-site digitization and interpretation, this approach holds promise for enhancing diagnostic reliability and efficiency across various clinical and public health settings.

## METHODS

### System Design and Processing Pipeline

The proposed AI-assisted system for LFA result reading, interpretation and digital reporting is presented in **Figure 1**. The digitization of LFAs was accomplished through a custom mobile application, TiraSpot (Spotlab, Madrid, Spain), that enabled users to capture consistent, high-quality images of the tests, taken between 2020-2024. The app’s camera interface displays a mask that matches the geometry and morphology of a specific LFA test, guiding the user to correctly align the LFA. This mask was later used as a reference for cropping the area of interest where the bands were located. Once the picture was taken, the app showed the image asking the user to confirm a correct acquisition (aligned with the mask and on focus), followed by a step where the user introduced the visual interpretation of the LFA result. Configuration of the app can be freely adapted to the specific needs of the LFA that is being digitized, by modifying either the mask or the possible outcomes. Moreover, images and related sample data were securely uploaded to a web platform, where images were accessed and reviewed, ensuring comprehensive oversight of the test outcomes.

**Figure 1.**
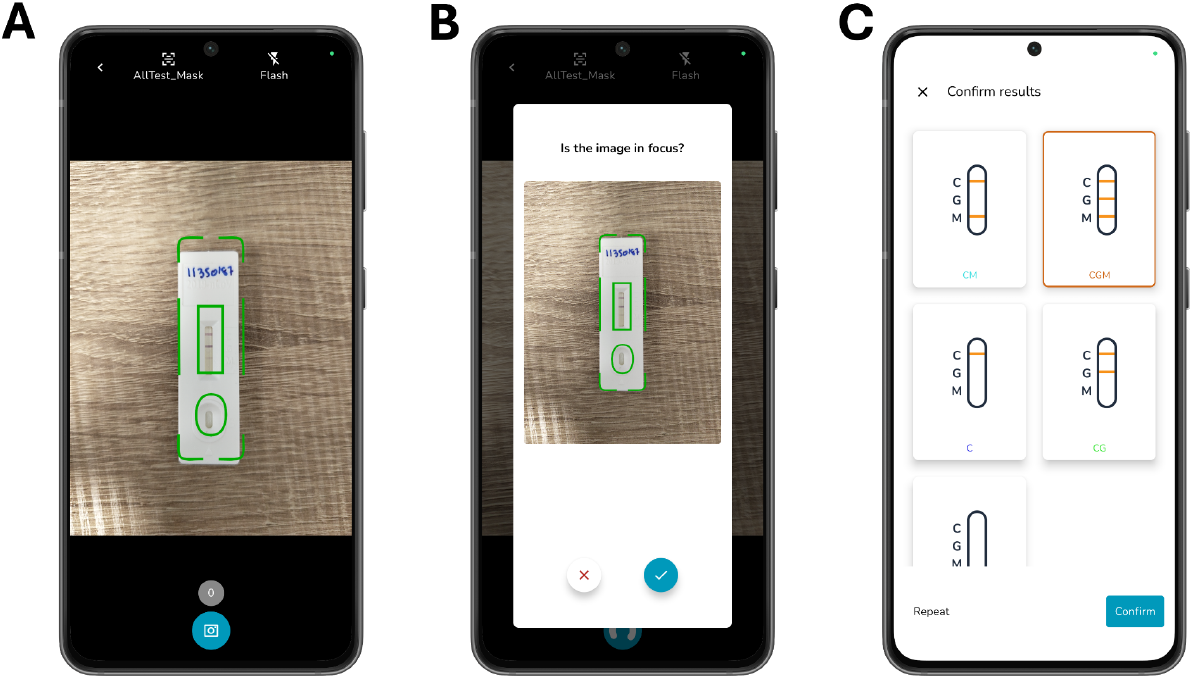
TiraSpot mobile app, showing (A) image capture interface for standardized LFA photographs, (B) image quality check screen to ensure the photo was in focus and properly aligned and (C) result confirmation interface allowing users to interpret and validate the LFA result.

### Artificial Intelligence Algorithm for Universal LFA Qualitative Reading

The entire image processing pipeline is described in **Figure 2**. The algorithm pre-processed the original photograph by cropping the image to extract only the area with the control and test bands. This new cropped image was downscaled to a smaller resolution of 325×100, and then pixel intensities were linearly scaled to the range [-1, 1] by applying min-max normalization. Finally, whitening was applied to decorrelate the pixels and ensure zero mean and unit variance. Synthetic augmentation techniques, such as changes in brightness or saturation, or random left and right flipping, were also employed during the training phase.

**Figure 2.**
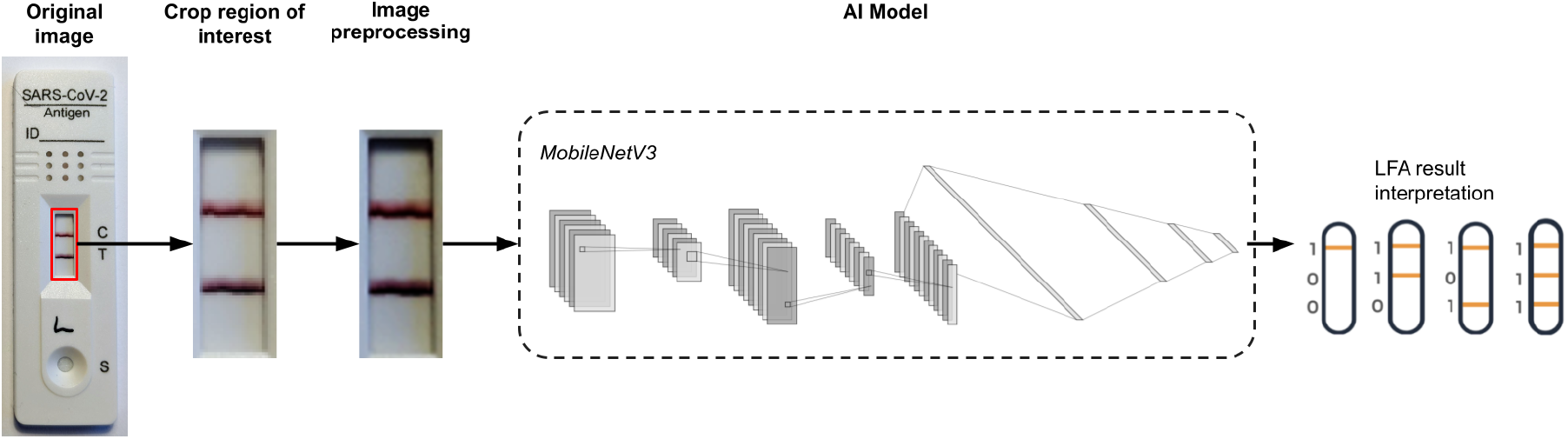
Image processing pipeline for LFA interpretation. The TiraSpot app captured an image of the test, which was then cropped to isolate the region of interest. The cropped image underwent preprocessing and was passed through an AI model to predict the LFA result. Visual results were translated into binary codes for AI analysis: for 2-band LFAs, valid outputs are limited to 100 and 110; for 3-band LFAs, possible valid results include 100, 110, 101, and 111.

The preprocessed image was introduced into a MobileNetV3 convolutional network *(25)* attached to a multi-layer perceptron. The final layer comprised 4 output neurons to predict the result, where each output corresponded to a specific binary code (e.g. 110, 110, 101, 111, see **Figure 2**), representing all possible combinations of band activations of the LFA. Here “1” indicated the presence of a signal for the respective band, while “0” indicated its absence. This structure accounts for up to three bands on an LFA, including one control and up to two test bands. This network architecture had been specifically designed to be lightweight so it could be efficiently integrated into smartphones.

To facilitate interpretation of individual band activations, a post-processing step was also employed. This step combined the probabilities of the relevant binary outputs to compute the likelihood of the presence for each band. This approach ensured accurate computation of band-specific probabilities.

### Data Acquisition and AI Performance Evaluation

To evaluate the system variability, each LFA was digitized and photographed, at least twice, by two or three different smartphone models with different technical capacities. To gain robustness and generalizability, more than 20 different smartphone models equipped with a ≥12 MPx camera were used in this study *(Supplementary Note 1)*, ranging from low-to high-range devices. In total, 17 types of LFAs were digitized, covering a range of diseases including malaria, cryptococcosis, COVID-19, and Chagas disease. These included two malaria LFAs, one cryptococcal antigen LFA, five COVID-19 LFAs (covering both antigen and antibody detection), and nine Chagas disease LFAs. A detailed list of the tests, including their manufacturers and abbreviations, is provided in **Table 1**, and example images for each of the test types are presented in **Figure 3**. All photographs form a dataset composed of 22,576 images, all corresponding to valid LFAs (i.e., tests with a visible control line). COVID-19 data were collected, processed, and accessed for research purposes between March and June 2021; Chagas disease data between January 2019 and March 2021; and malaria data between February and July 2024. No human samples were used for the cryptococcosis dataset. The authors did not have access to potentially identifying information during or after data collection.

**Table 1.** Detailed description of the types of LFAs digitized using the TiraSpot system and included in the study.

| Disease | Test Name | Manufacturer | Abbreviation |
| --- | --- | --- | --- |
| Malaria | BinaxNOW,™ Malaria | Abbott | MAL-BINAX |
| Malaria | Bioline,™ Malaria Ag P.f/Pan | Abbott | MAL-BIOLINE |
| Cryptococcosis | Cryptococcal Antigen LFA CrAg LFA | IMMY | CRAG-QL |
| COVID-19 | 2019-nCoV IgG/IgM Rapid Test Cassette | Hangzhou AllTest Biotech Co. Ltd. | COVID-ALL |
| COVID-19 | Panbio COVID-19 IgG/IgM Rapid Test Device | Abbott | COVID-ABB |
| COVID-19 | UNscience COVID-19 IgG/IgM Rapid Test | Wuhan UNscience Biotechnology Co. Ltd. | COVID-SCI |
| COVID-19 | SARS-CoV-2 Rapid Antibody Test | Roche | COVID-ROCHE |
| COVID-19 | Panbio COVID-19 Ag Rapid Test Device | Abbott | COVID-ABBAG |
| Chagas | Chagas Ad-Bio Combo Rapid Test | CTK Biotech | CH-ADBIO |
| Chagas | Chagas Ab Rapid Test | Artron | CH-ARTR |
| Chagas | Chagas Rapido First Response | Lemos Lab | CH-FIRST |
| Chagas | Hexagon Chagas | Human | CH-HEX |
| Chagas | Chagas Detect Plus Rapid Test | InBios Inc. | CH-PLUS |
| Chagas | SD Chagas Ab Rapid | Standard Diagnostic | CH-SDAB |
| Chagas | Chagas Stat-Pak Assay | Chembio | CH-STAT |
| Chagas | WL Check Chagas | Wiener Lab | CH-WLCHECK |
| Chagas | Chagas Ab Xerion Cassette | Xerion | CH-XERION |

**Figure 3.**
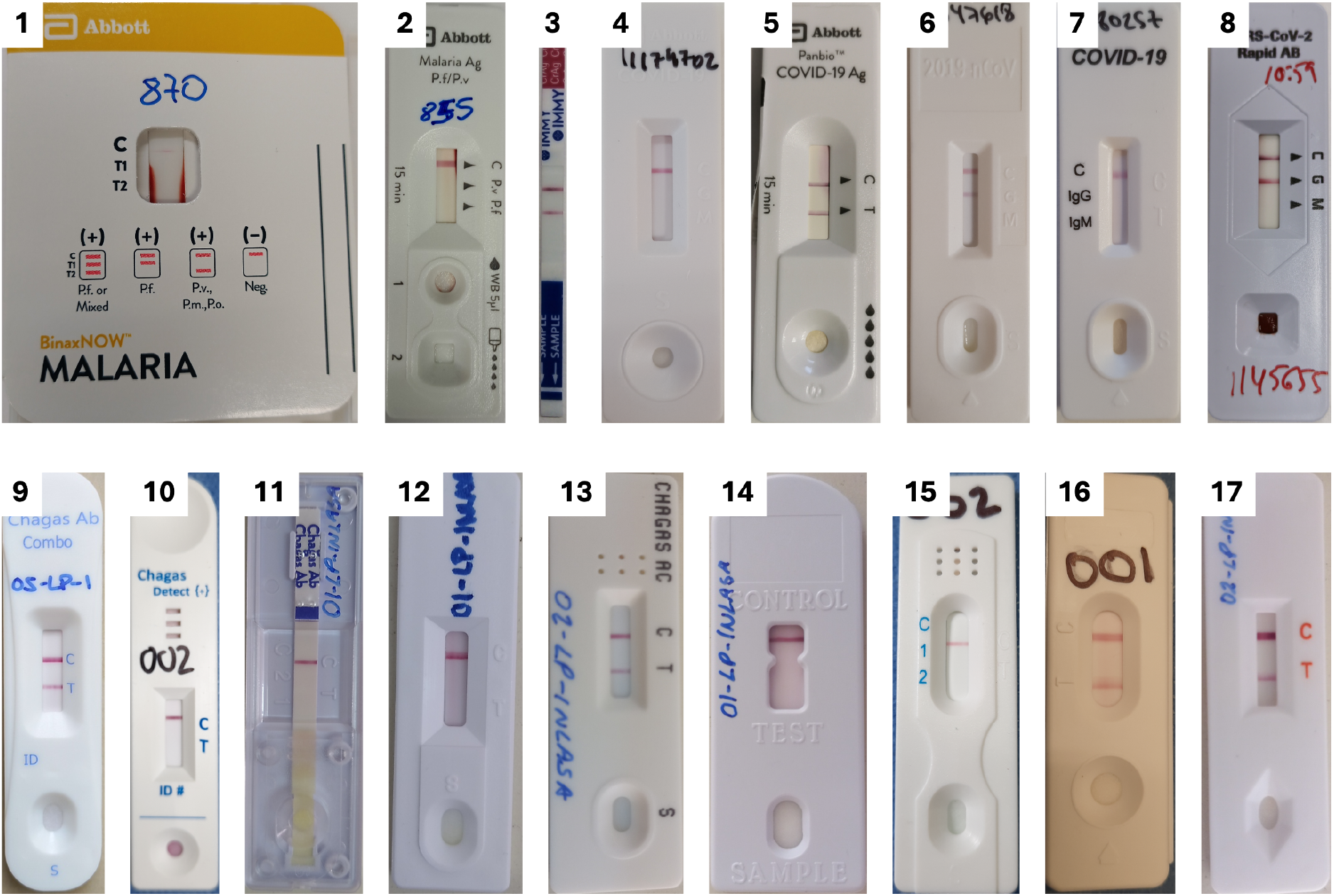
Visualization of the 17 different types of LFAs used in this study. MAL-BINAX (1), MAL-BIOLINE (2), CRAG-QL (3), COVID-ABB (4), COVID-ABBAG (5), COVID-ALL (6), COVID-SCI (7), COVID-ROCHE (8), CH-ADBIO (9), CH-FIRST (10), CH-SDAB (11), CH-WLCHECK (12), CH-XERION (13), CH-STAT (14), CH-ARTR (15), CH-HEX (16) and CH-PLUS (17).

To generate the ground-truth, each LFA was visually interpreted by, at least, two different observers (health care professionals, experts on the specific disease diagnosis and trained for results interpretation of those specific LFAs) and classified as positive or negative for the specific test bands. In cases of discrepancy between observers, a third one decides which was the final result after a careful reading of the questioned picture. Binary results (positive or negative) were assigned for each band based on visual interpretation. The image dataset per LFA, band and visual result is summarized in Table 2.

**Table 2.**
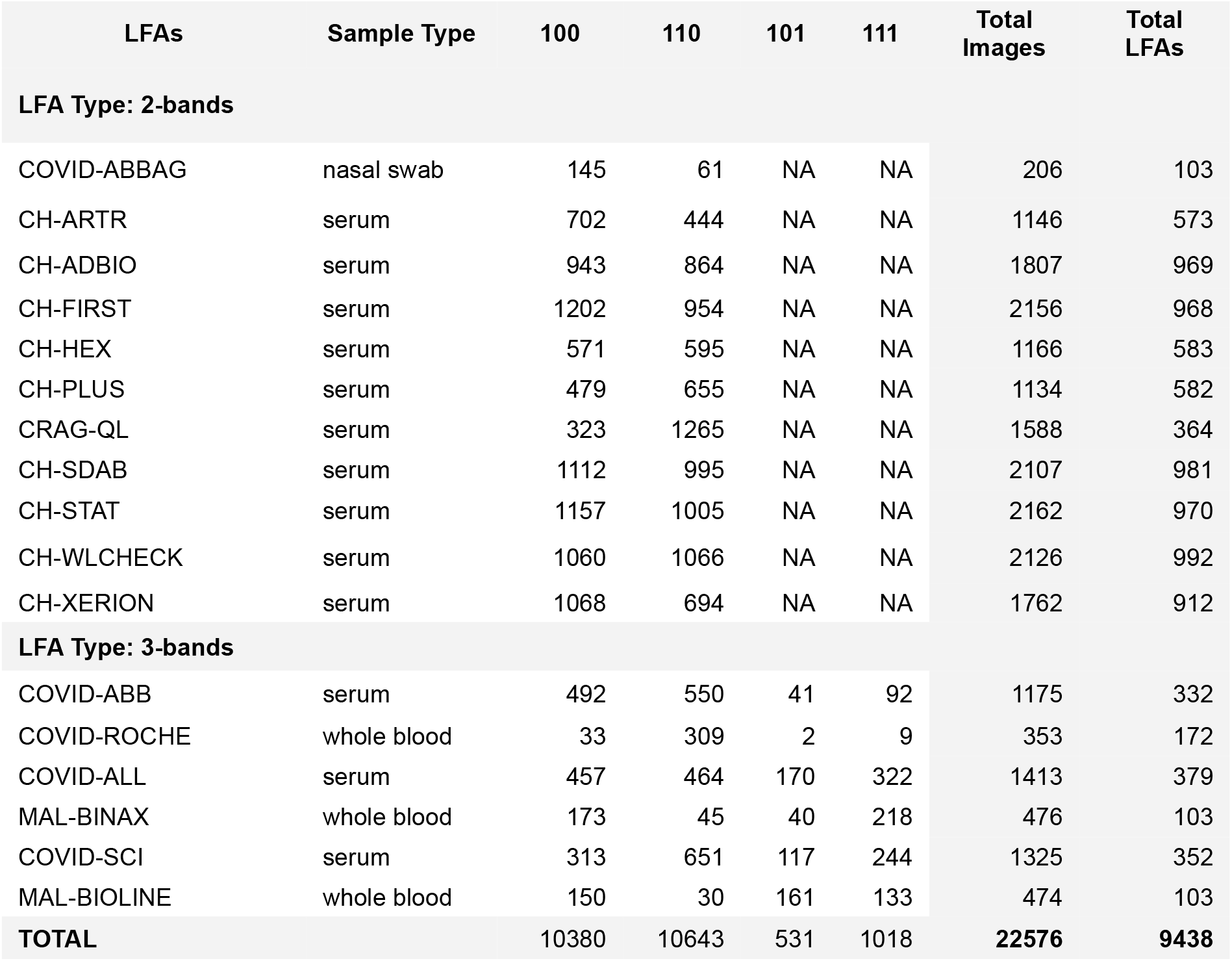
Image data distribution per Lateral Flow Assay (LFA) brand types, categorized by sample type and binary result (100, 110, 101, 111). See Figure 2 for binary result visual translation. NA, not applicable.

To validate the AI model’s performance, a leave-one-out approach was employed. In each experiment, all data from one LFA type was withheld from the training set, ensuring the model had no prior exposure to that type. A test set of 100 images from the withheld type were used to evaluate the model’s ability to generalize to unseen LFA types. During the test set construction, a maximum of one image per specific LFA was allowed, ensuring no repeated images for a single physical LFA unit. This process was repeated 10 times for each LFA type, resulting in 10 independent models and 1000 test inferences, which were evaluated after a bootstrapping process of 1000 iterations with replacement.

In later experiments, incremental samples of 10, 30, 40, and 50 images of the withheld LFA type were included in the training set. This allowed for monitoring the model’s ability to learn from a small number of examples of a specific LFA type, in contrast to the leave-one-out experiment where the model was entirely unexposed to the withheld LFA type for training. As in the previous phase, this process was repeated 10 times for each LFA type, with independently sampled test and training sets in each iteration, and an evaluation process based on bootstrapping with replacement.

This iterative process allowed for a rigorous assessment of the models accuracy and generalization capabilities across various LFA types. Performance of the AI algorithm for qualitative test result interpretation was assessed in terms of the area under the ROC curve (AUC), sensitivity (SN), specificity (SP), and the overall accuracy (ACC) for each band of the LFA in the validation dataset. The Youden index was used for threshold selection when determining a label-wise positive or negative outcome by the model, as it maximizes the balance between SN and SP.

### Ethical considerations

This study adhered to the Declaration of Helsinki. COVID-19 samples were approved by the Clinical Research Ethics Committee of Ramón y Cajal University Hospital (Ref: 127/21), and malaria samples by the Ethics Committee of the Instituto de Salud Carlos III (Ref: CEI PI 74_2020). For Chagas disease, 555 participants provided written informed consent; 30 additional anonymized residual blood-donor samples from INS quality control were used under Colombian regulations. Data were coded and handled confidentially (Law 23/1981, Article 34), and the Chagas-related work was considered minimal risk (Resolution 8430, 1993). For cryptococcosis, IRB review and informed consent were not applicable as no human experiments were performed.

## RESULTS

### Performance of the Universal AI Model for LFA Interpretation

The model’s performance was assessed under a leave-one-LFA-type-out evaluation to test its generalization capability. The following results summarize the model’s performance under this evaluation framework.

The proposed universal AI model for LFA result interpretation demonstrated strong accuracy and reliability for interpreting a wide range of LFAs across different settings. **Table 3** summarizes the performance metrics per LFA type and band when comparing AI predictions and visual interpretations of the LFAs (used as ground truth), with a mean AUC of 94,3 for the detection of the second band, and a mean AUC of 83,9 for the third band. Sensitivity and specificity values consistently exceeded 90% for the second band detection across most test types, with certain exceptions in low-signal scenarios. This is the case of MAL-BIOLINE LFAs, where background interference occasionally led to false positives, resulting in lower specificity metrics as compared to other LFAs.

**Table 3.**
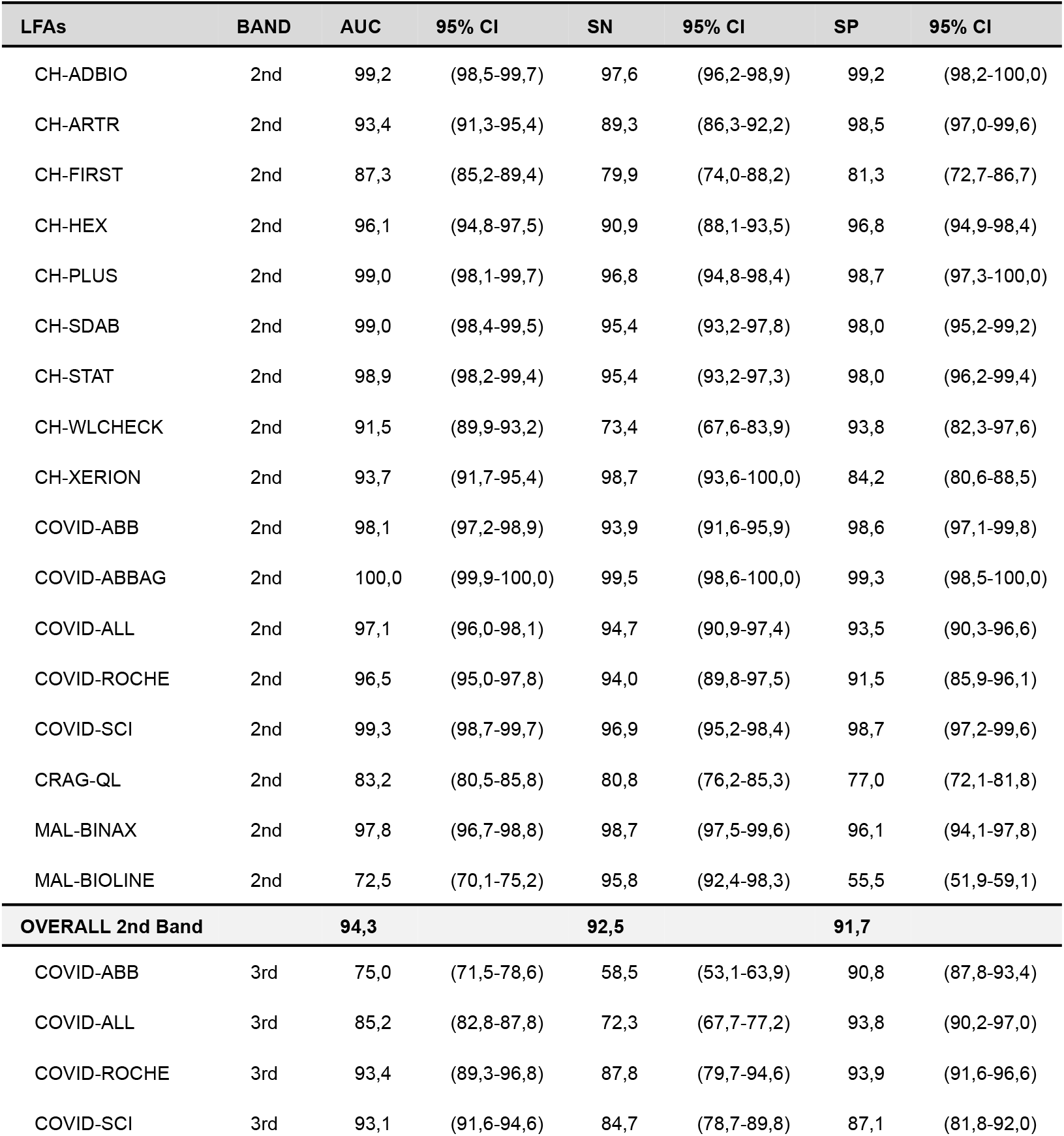
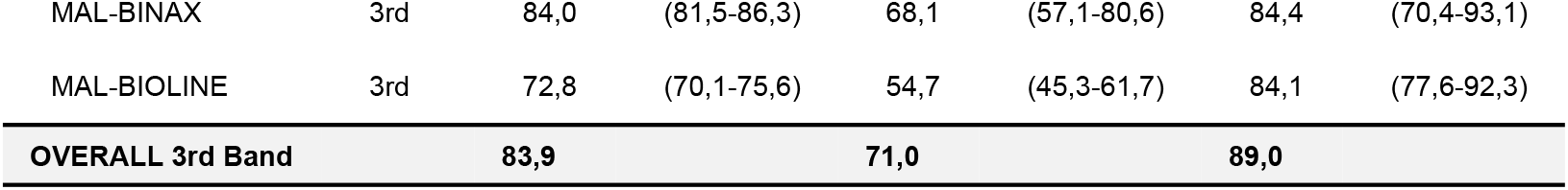
Performance of the artificial intelligence algorithm for predicting LFA results with respect to human visual reading by test type and test band, with zero training samples for the measured test type. *CI, Confidence Interval; AUC, Area under the curve; SN and SP, Sensitivity and Specificity*.

| LFAs | BAND | AUC | 95% CI | SN | 95% CI | SP | 95% CI |
| --- | --- | --- | --- | --- | --- | --- | --- |
| CH-ADBIO | 2nd | 99,2 | (98,5-99,7) | 97,6 | (96,2-98,9) | 99,2 | (98,2-100,0) |
| CH-ARTR | 2nd | 93,4 | (91,3-95,4) | 89,3 | (86,3-92,2) | 98,5 | (97,0-99,6) |
| CH-FIRST | 2nd | 87,3 | (85,2-89,4) | 79,9 | (74,0-88,2) | 81,3 | (72,7-86,7) |
| CH-HEX | 2nd | 96,1 | (94,8-97,5) | 90,9 | (88,1-93,5) | 96,8 | (94,9-98,4) |
| CH-PLUS | 2nd | 99,0 | (98,1-99,7) | 96,8 | (94,8-98,4) | 98,7 | (97,3-100,0) |
| CH-SDAB | 2nd | 99,0 | (98,4-99,5) | 95,4 | (93,2-97,8) | 98,0 | (95,2-99,2) |
| CH-STAT | 2nd | 98,9 | (98,2-99,4) | 95,4 | (93,2-97,3) | 98,0 | (96,2-99,4) |
| CH-WLCHECK | 2nd | 91,5 | (89,9-93,2) | 73,4 | (67,6-83,9) | 93,8 | (82,3-97,6) |
| CH-XERION | 2nd | 93,7 | (91,7-95,4) | 98,7 | (93,6-100,0) | 84,2 | (80,6-88,5) |
| COVID-ABB | 2nd | 98,1 | (97,2-98,9) | 93,9 | (91,6-95,9) | 98,6 | (97,1-99,8) |
| COVID-ABBAG | 2nd | 100,0 | (99,9-100,0) | 99,5 | (98,6-100,0) | 99,3 | (98,5-100,0) |
| COVID-ALL | 2nd | 97,1 | (96,0-98,1) | 94,7 | (90,9-97,4) | 93,5 | (90,3-96,6) |
| COVID-ROCHE | 2nd | 96,5 | (95,0-97,8) | 94,0 | (89,8-97,5) | 91,5 | (85,9-96,1) |
| COVID-SCI | 2nd | 99,3 | (98,7-99,7) | 96,9 | (95,2-98,4) | 98,7 | (97,2-99,6) |
| CRAG-QL | 2nd | 83,2 | (80,5-85,8) | 80,8 | (76,2-85,3) | 77,0 | (72,1-81,8) |
| MAL-BINAX | 2nd | 97,8 | (96,7-98,8) | 98,7 | (97,5-99,6) | 96,1 | (94,1-97,8) |
| MAL-BIOLINE | 2nd | 72,5 | (70,1-75,2) | 95,8 | (92,4-98,3) | 55,5 | (51,9-59,1) |
| **OVERALL 2nd Band** |  | **94,3** |  | **92,5** |  | **91,7** |  |
| COVID-ABB | 3rd | 75,0 | (71,5-78,6) | 58,5 | (53,1-63,9) | 90,8 | (87,8-93,4) |
| COVID-ALL | 3rd | 85,2 | (82,8-87,8) | 72,3 | (67,7-77,2) | 93,8 | (90,2-97,0) |
| COVID-ROCHE | 3rd | 93,4 | (89,3-96,8) | 87,8 | (79,7-94,6) | 93,9 | (91,6-96,6) |
| COVID-SCI | 3rd | 93,1 | (91,6-94,6) | 84,7 | (78,7-89,8) | 87,1 | (81,8-92,0) |
| MAL-BINAX | 3rd | 84,0 | (81,5-86,3) | 68,1 | (57,1-80,6) | 84,4 | (70,4-93,1) |
| MAL-BIOLINE | 3rd | 72,8 | (70,1-75,6) | 54,7 | (45,3-61,7) | 84,1 | (77,6-92,3) |
| **OVERALL 3rd Band** |  | **83,9** |  | **71,0** |  | **89,0** |  |

For the third band, mean sensitivity and specificity were 71% and 89% respectively. This lower performance compared to the second band is probably due to the imbalance of the dataset, where 3-band tests are significantly underrepresented among the LFA types. Also, and with the exception of MAL-BIOLINE, positive examples for the third band are underrepresented among the 3-band LFAs (**Table 2**). Furthermore, the third bands in nearly all 3-band LFAs, particularly the COVID-19 tests, consistently exhibited very faint signals that were almost invisible in the photographs, further contributing to the reduced performance. These findings emphasize that additional training data may be beneficial for improving accuracy in challenging test types.

Examples of cases showing result discrepancies in problematic LFA types are represented in **Figure 4**. Background, band intensity and sample type affected model performance. COVID-ABB (**Figure 4, 1-4**) and COVID-ALL LFA types (**Figure 4, 5-8**) often showed very faint positive bands leading to failures in third band detections, which explains the low sensitivity (**Table 3**). In LFAs that utilize whole-blood samples, partially coagulated blood can hinder strip absorption and background clearance. This issue is evident in the MAL-BINAX (**Figure 4, 9-12**) and MAL-BIOLINE (**Figure 4, 13-16**) malaria-antigen detection LFAs, where many strip images displayed intense backgrounds or sample stains, resulting in lower performance metrics compared to other LFAs that typically use plasma and serum samples. Specifically in the case of MAL-BINAX, sample stains tend to adhere to the bottom of the strip, causing problems with the third band, while the second band maintains relatively good performance (**Table 3**).

**Figure 4.**
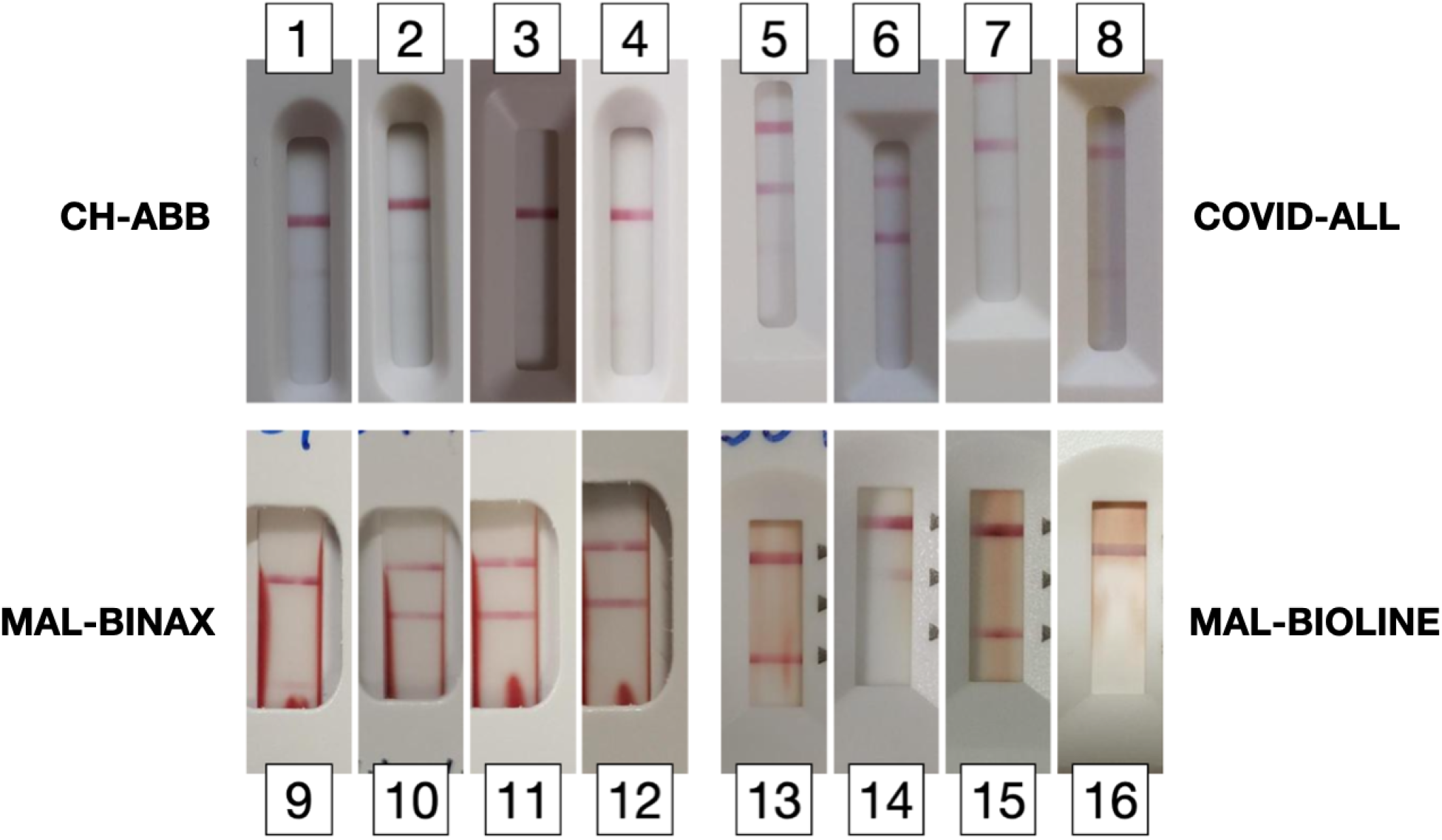
Examples of LFA cases showing discrepancies in third band detections. In the COVID-ABB set (images 1 to 4), visual interpretation results (binary result) are, from left to right, 110, 110, 101 and 101. In the COVID-ALL set (images 5 to 8), visual interpretation results are 111, 111, 111 and 101. In the MAL-BINAX set (images 9 to 12), visual interpretation results are 101, 110, 110 and 110. In the MAL-BIOLINE set (images 13 to 16), visual interpretation results are 101, 110, 101 and 100.

**Figure 5.**
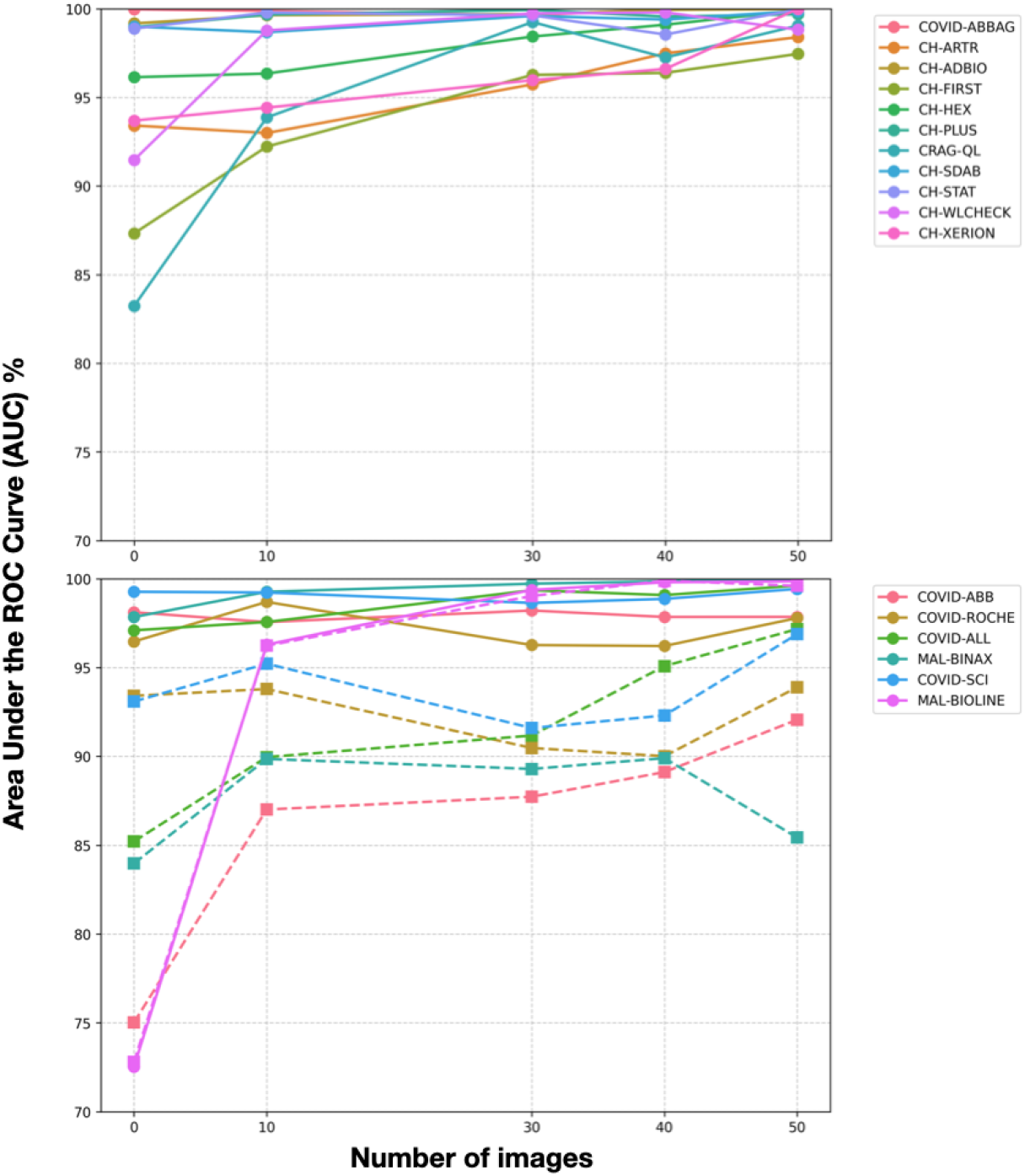
Performance evolution through model training with incremental number of LFA images, for 2 band LFAs (above) and 3 band LFAs (below). Metrics for second band results are represented as a solid-line, while metrics for third band results are represented as a dotted-line

### Incremental Training Results

Figure 5 shows the model’s performance improvement as additional examples of a given LFA type are introduced into the training set. This progressive improvement underscores the scalability and adaptability of the AI system for universal LFA interpretation.

For the second-band detection, the overall AUC improved from 94,3% in the leave-one-out experiment (with no LFA-specific images used in training) to 98,5% and 99,3% as the training dataset expanded to include 30 and 50 LFA-specific images (Supplementary Table 1 and 2), respectively. This trend highlights the ability of the model to achieve substantial accuracy gains with relatively modest increases in training data. Most test models demonstrated robust performance after incorporating 30 to 50 LFA-specific images, achieving an AUC over 99% across most 2-band LFAs and over 97% for the second band in 3-band LFAs.

The performance for the third band predictions in 3-band LFAs was consistently lower, requiring a greater number of images for comparable accuracy. These results reflect the model’s ability to improve its accuracy even in challenging scenarios, such as limited training data or very faint band signals during incremental training. Initially, in the leave-one-out scenario, the overall AUC for the third band was 83,9%, improving to 91,6% and 94,2% with the inclusion of 30 and 50 specific-LFA images (Supplementary Table 1 and 2), respectively. In the case of MAL-BIOLINE, where the number of positive examples for the third band is significantly higher than in other LFA types (**Table 2**), the background intensity problem associated with the sample type is overcome with only a few training examples, approaching an AUC of 100% (panel B pink line). However, in the case of MAL-BINAX LFA, the lower performance persists even when training with incremental image samples. This can be attributed to the particular cardboard-based LFA design, which may impact sample proper absorption and clear backgrounds strips. These results suggest that band-specific image data imbalance and test design complexity may play a crucial role in AI model performance.

## DISCUSSION

We developed and validated a novel AI model for universal reading and interpretation of lateral flow assays (LFAs) that demonstrate robust performance across diverse test types without the need for additional hardware. The system operates through a smartphone-based mobile application that standardized LFA image capture, and performs reliably on mid-range devices equipped with standard cameras (≥12Mpx). This accessibility makes the approach broadly deployable across different platforms and healthcare settings.

The AI models achieve an overall AUC of 91,6% (Sensitivity=86,9%, Specificity=91,0%) for previously unseen LFA models, with significant improvements to 98% (Sensitivity=97,1%, Specificity=95,4%) after incorporating 50 LFA specific images for training, respectively. This result underscores the model’s ability to rapidly adapt to new LFAs with minimal additional data, demonstrating that even if the performance for a specific LFA is initially suboptimal, adding a few representative images can significantly improve accuracy. The AI model was trained and validated on a dataset encompassing 17 different LFA types and 22,576 images, including assays for SARS-CoV-2, malaria, Chagas, and Cryptococcal infections. This diverse training set enabled the development of versatile models capable of identifying and interpreting multiple band configurations.

Our AI system detects individual test bands, enabling multi-band recognition and different signal intensities for different analytes, allowing a broader use than previous approaches that were limited to specific test types or simpler band patterns. The proposed AI model works well in 2-band detection scenarios, achieving more than 96% AUC across most test configurations. However, the model’s performance on 3-band LFAs revealed opportunities for improvement (overall AUC of 88,7%, including both second and third band identification), primarily due to a limited training dataset with imbalance regarding positive examples for the third band.

A key advantage of this system is its hardware-free operation, eliminating the need for adapters, proprietary cassettes, or controlled lighting. By using an app that normalizes the way images are captured with smartphones, it reduces costs, improves scalability, and could enable deployment in resource-limited settings, making it a potential reasonable choice for point-of-care diagnostics and mobile health clinics. Moreover, the AI model can be fully integrated into the mobile app for real-time inference within milliseconds (and without requiring an internet connection), ensuring instant LFA result interpretation to assist healthcare professionals in decision-making. Additionally, the system can also be integrated into a cloud platform, allowing for real-time epidemiological surveillance and public health monitoring, facilitating rapid outbreak detection and improved disease tracking, as already applied by different research groups in different settings (16,26,27).

According to the most recent TPPs outlined by WHO and FIND Diagnostics (8–10), AI models for LFA interpretation should maintain high accuracy under various conditions while remaining computationally efficient. Our AI model was designed to meet these requirements, achieving the mandated minimum 95% accuracy with just 30 to 50 training images of the specific LFA type being tested, while enabling deployment on resource-constrained devices.

Future iterations of the AI system could explore enhanced capabilities for quantitative estimation of analytes through LFA band intensity determination, as previously described for the Cryptococcal antigen LFA (21,22). From a practical and implementation perspective, a universal reader allows using the same app to read and report multiple LFAs done to a single patient. It can also adapt the tests available to the user based, for instance, on GPS coordinates. Health Systems could implement a single system for LFA reading which incorporates different brands of tests as they are approved for use-kind of an “LFA wallet”.

In conclusion, this study represents a new methodological strategy and a simple tool for automated mobile LFA interpretation, demonstrating robust performance across images from multiple LFAs under varying lighting conditions using multiple smartphone models with different technical and camera capabilities. By addressing subjectivity in visual interpretation, enabling real-time digital reporting, and supporting epidemiological surveillance, this system can contribute to enhancing diagnostic accessibility and disease monitoring worldwide.

## Data Availability

All data produced in the present study are available upon reasonable request to the authors.

## SUPPORTING INFORMATION

**S1 Note**. List of smartphone models used for the acquisition of the LFA images.

| Smartphone model | Rear camera |
| --- | --- |
| Samsung Galaxy A33 5G | 48 MP · f/1.8 |
| OPPO Reno6 5G | 64 MP · f/1.7 |
| Samsung Galaxy S9+ | 12 MP · f/1.5–2.4 |
| Motorola Moto E6 Play | 13 MP · f/2.2 |
| Samsung Galaxy S9 | 12 MP · f/1.5–2.4 |
| Samsung Galaxy S10+ | 12 MP · f/1.5–2.4 |
| Xiaomi Pocophone F1 | 12 MP · f/1.9 |
| Samsung Galaxy A51 | 48 MP · f/2.0 |
| Xiaomi Redmi Note 8T | 48 MP · f/1.8 |
| BQ Aquaris X2 | 12 MP · f/1.8 |
| Huawei P30 | 40 MP · f/1.8 |
| Xiaomi Redmi Note 7 | 48 MP · f/1.8 |
| Xiaomi Mi 9T | 48 MP · f/1.75 |
| Apple iPhone XR | 12 MP · f/1.8 |
| Samsung Galaxy S21 | 12 MP · f/1.8 |
| Google Pixel 6 Pro | 50 MP · f/1.85 |
| Xiaomi Redmi Note 10 (5G) | 48 MP · f/1.8 |
| Xiaomi POCO X4 GT | 64 MP · f/1.9 |
| OnePlus 5 | 16 MP · f/1.7 |
| Samsung Galaxy A32 | 64 MP · f/1.8 |
| Samsung Galaxy A31 | 48 MP · f/2.0 |
| LG K22 | 13 MP · f/2.2 |

**S1 Table.**
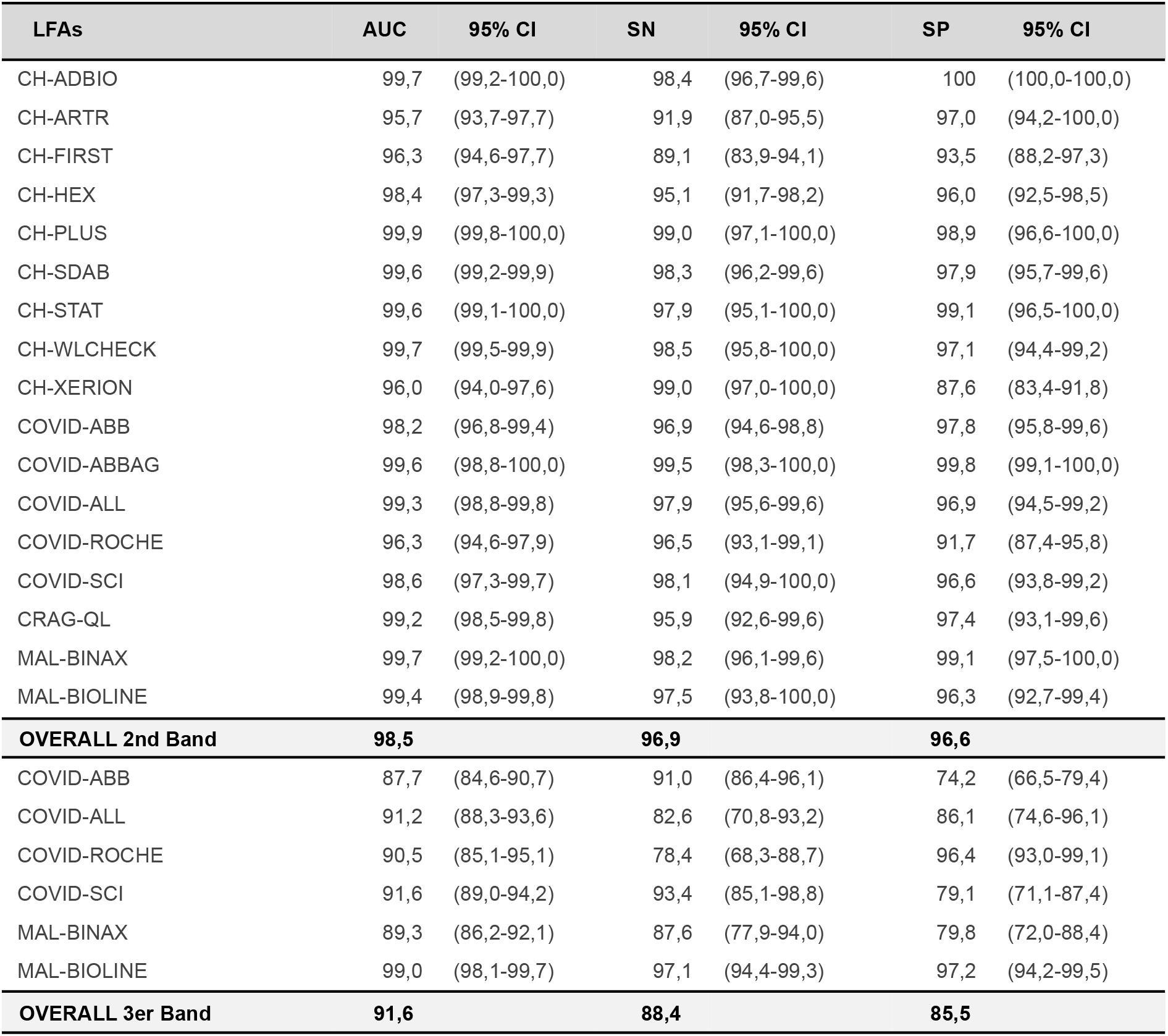
Performance of the artificial intelligence algorithm for predicting LFA results with respect to human visual reading by test type and test band, with 30 training samples for the measured test type. *CI, Confidence Interval; AUC, Area under the curve; SN and SP, Sensitivity and Specificity*.

| LFA | AUC | 95% CI | SN | 95% CI | SP | 95% CI |
| --- | --- | --- | --- | --- | --- | --- |
| CH-ADBIO | 99,7 | (99,2-100,0) | 98,4 | (96,7-99,6) | 100 | (100,0-100,0) |
| CH-ARTR | 95,7 | (93,7-97,7) | 91,9 | (87,0-95,5) | 97,0 | (94,2-100,0) |
| CH-FIRST | 96,3 | (94,6-97,7) | 89,1 | (83,9-94,1) | 93,5 | (88,2-97,3) |
| CH-HEX | 98,4 | (97,3-99,3) | 95,1 | (91,7-98,2) | 96,0 | (92,5-98,5) |
| CH-PLUS | 99,9 | (99,8-100,0) | 99,0 | (97,1-100,0) | 98,9 | (96,6-100,0) |
| CH-SDAB | 99,6 | (99,2-99,9) | 98,3 | (96,2-99,6) | 97,9 | (95,7-99,6) |
| CH-STAT | 99,6 | (99,1-100,0) | 97,9 | (95,1-100,0) | 99,1 | (96,5-100,0) |
| CH-WLCHECK | 99,7 | (99,5-99,9) | 98,5 | (95,8-100,0) | 97,1 | (94,4-99,2) |
| CH-XERION | 96,0 | (94,0-97,6) | 99,0 | (97,0-100,0) | 87,6 | (83,4-91,8) |
| COVID-ABB | 98,2 | (96,8-99,4) | 96,9 | (94,6-98,8) | 97,8 | (95,8-99,6) |
| COVID-ABBAG | 99,6 | (98,8-100,0) | 99,5 | (98,3-100,0) | 99,8 | (99,1-100,0) |
| COVID-ALL | 99,3 | (98,8-99,8) | 97,9 | (95,6-99,6) | 96,9 | (94,5-99,2) |
| COVID-ROCHE | 96,3 | (94,6-97,9) | 96,5 | (93,1-99,1) | 91,7 | (87,4-95,8) |
| COVID-SCI | 98,6 | (97,3-99,7) | 98,1 | (94,9-100,0) | 96,6 | (93,8-99,2) |
| CRAG-QL | 99,2 | (98,5-99,8) | 95,9 | (92,6-99,6) | 97,4 | (93,1-99,6) |
| MAL-BINAX | 99,7 | (99,2-100,0) | 98,2 | (96,1-99,6) | 99,1 | (97,5-100,0) |
| MAL-BIOLINE | 99,4 | (98,9-99,8) | 97,5 | (93,8-100,0) | 96,3 | (92,7-99,4) |
| **OVERALL 2nd Band** | **98,5** |  | **96,9** |  | **96,6** |  |
| COVID-ABB | 87,7 | (84,6-90,7) | 91,0 | (86,4-96,1) | 74,2 | (66,5-79,4) |
| COVID-ALL | 91,2 | (88,3-93,6) | 82,6 | (70,8-93,2) | 86,1 | (74,6-96,1) |
| COVID-ROCHE | 90,5 | (85,1-95,1) | 78,4 | (68,3-88,7) | 96,4 | (93,0-99,1) |
| COVID-SCI | 91,6 | (89,0-94,2) | 93,4 | (85,1-98,8) | 79,1 | (71,1-87,4) |
| MAL-BINAX | 89,3 | (86,2-92,1) | 87,6 | (77,9-94,0) | 79,8 | (72,0-88,4) |
| MAL-BIOLINE | 99,0 | (98,1-99,7) | 97,1 | (94,4-99,3) | 97,2 | (94,2-99,5) |
| **OVERALL 3er Band** | **91,6** |  | **88,4** |  | **85,5** |  |

**S2 Table.**
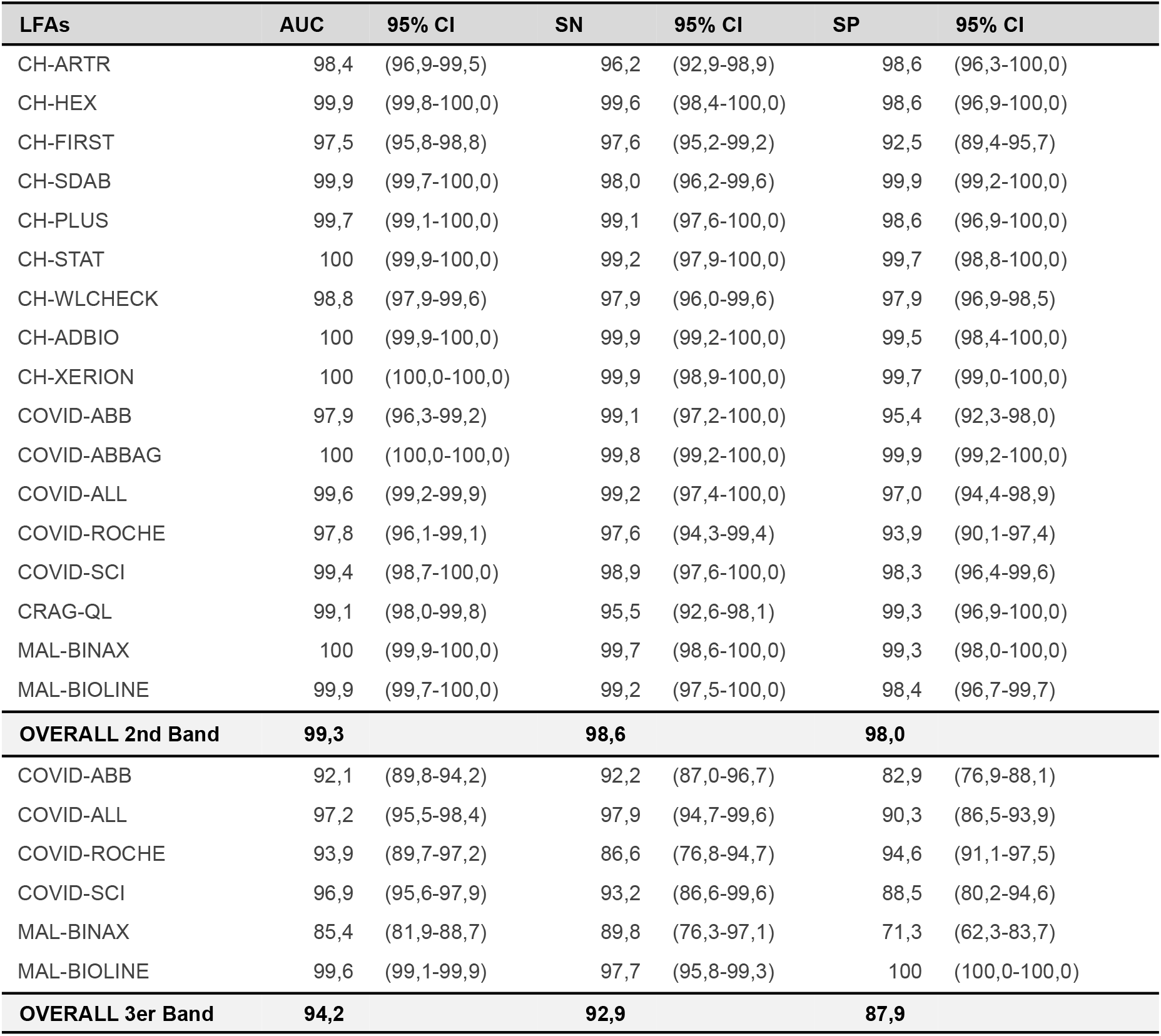
Performance of the artificial intelligence algorithm for predicting LFA results with respect to human visual reading by test type and test band, with 50 training samples for the measured test type. *CI, Confidence Interval; AUC, Area under the curve; SN and SP, Sensitivity and Specificity*.

